# Determinants of Adequate Antenatal Care Attendance in Nigeria: A Survey-Weighted Analysis of the 2018 Demographic and Health Survey

**DOI:** 10.64898/2026.05.02.26352203

**Authors:** Ugoeze Lucy Unegbu

**Author notes:** **Funding:** This study did not receive any funding. **Data availability:** Data are publicly available upon registration at dhsprogram.com.

## Abstract

**Background:** Nigeria accounts for approximately 19% of global maternal deaths, yet 4 in 10 Nigerian women do not meet the World Health Organization minimum standard of four antenatal care (ANC) visits. Understanding which women are being left behind, and how early they initiate care, is essential for designing effective maternal health programmes.

**Methods:** We conducted a cross-sectional analysis of 21,465 women with a birth in the five years preceding the 2018 Nigeria Demographic and Health Survey (NDHS). Survey-weighted multivariable logistic regression was used to estimate adjusted odds ratios (aOR) for seven sociodemographic predictors of adequate ANC attendance (4 or more visits). Kaplan-Meier survival analysis and Cox proportional hazards modelling were additionally applied to 16,084 women with complete ANC timing data to examine time to first ANC visit. Confounding was quantified by comparing crude and adjusted estimates.

**Results:** The national weighted prevalence of adequate ANC was 57.8% (95% CI: 56.2%-59.4%). The median gestational age at first ANC visit was 5 months, two months later than WHO recommendations. Higher education (aOR = 5.64, 95% CI: 4.45-7.15) and richest wealth quintile (aOR = 3.93, 95% CI: 3.11-4.95) were the strongest independent predictors. Urban residence lost significance entirely after adjustment (aOR = 1.12, p = 0.113), indicating that the crude urban advantage is fully explained by the higher education and wealth of urban women. Educated women initiated ANC 35% faster than uneducated women (HR = 1.35, 95% CI: 1.23-1.47). Confounding was substantial: 74.9% of higher education’s crude effect was attributable to correlated socioeconomic factors.

**Conclusions:** Education and wealth are the dominant independent determinants of both adequate ANC attendance and earlier ANC initiation in Nigeria. The apparent urban advantage is entirely confounded. Targeted investment in girls’ education, wealth-sensitive demand-side financing, and community-based early ANC outreach particularly in the North West and North East are urgently needed.

## 1. Introduction

Maternal mortality remains a defining global health challenge, with approximately 295,000 maternal deaths occurring annually, the vast majority in low- and middle-income countries (LMICs) (World Health Organization [WHO], 2019). Nigeria bears a disproportionate share of this burden, accounting for approximately 19% of all global maternal deaths, with an estimated 512 deaths per 100,000 live births in 2017, among the highest rates recorded worldwide (WHO, 2019). Most of these deaths are attributable to direct obstetric complications including hemorrhage, hypertensive disorders, sepsis, and obstructed labor, conditions that are largely preventable or manageable with timely, adequate antenatal and intrapartum care (Say et al., 2014). Achieving the Sustainable Development Goal target of reducing the global maternal mortality ratio to fewer than 70 per 100,000 live births by 2030 will require substantial improvements in ANC coverage, particularly in high-burden settings such as Nigeria (United Nations, 2015).

Antenatal care provides a critical platform for delivering evidence-based interventions during pregnancy. These include blood pressure monitoring for pre-eclampsia detection, malaria prophylaxis through intermittent preventive treatment in pregnancy (a priority in Nigeria’s endemic setting), iron and folic acid supplementation, HIV testing and prevention of mother-to-child transmission counselling, foetal growth monitoring, and birth preparedness education (WHO, 2002). The timing of ANC initiation is as important as the total number of visits: a woman who attends her first visit at seven months of pregnancy has a compressed window of approximately six to eight weeks in which to receive these interventions, compared with seven months for a woman who initiates care in the second month. Late initiation therefore not only reduces the number of visits achievable before delivery but also limits the clinical benefit of each visit.

The WHO 2002 standard of a minimum of four ANC visits remains the reference standard captured in the 2018 Nigeria Demographic and Health Survey (NDHS) and forms the basis of the outcome measure used in this analysis. The 2018 NDHS reported a national ANC coverage of 57.8%, concealing extreme geographic and socioeconomic heterogeneity. In the North West geopolitical zone, adequate ANC coverage was 42.3%, compared with 89.9% in the South West a gap of nearly 48 percentage points within a single country. These disparities reflect the clustering of structural disadvantage low educational attainment, poverty, and rural residence in the same populations least likely to access care.

A substantial body of literature has identified education, wealth, urban residence, and parity as correlates of ANC utilization in sub-Saharan African settings (Fagbamigbe & Idemudia, 2015; Tessema, Z. T., et. al 2021). However, since these factors are highly correlated with each other, crude associations substantially overestimate the independent contribution of any single predictor. Studies that do not adjust for confounding risk misattributing the effect of wealth to education, or the effect of education to urban residence therefore leading to inefficiently targeted policies. Furthermore, most existing analyses examine only whether women attend adequate ANC, not when they initiate care. The timing dimension is clinically critical yet rarely modelled in the Nigerian literature.

This study addresses these gaps using data from the 2018 NDHS with two complementary analytic approaches: survey-weighted multivariable logistic regression to identify independent predictors of adequate ANC attendance, and Kaplan-Meier survival analysis with Cox proportional hazards modelling to examine the timing of ANC initiation and its independent determinants. This paper is the direct scientific sequel to a companion analysis of skilled birth attendance (SBA) using the same dataset, which found that attending four or more ANC visits was the strongest modifiable predictor of SBA (aOR = 3.80, 95% CI: 3.51-4.11) more influential than education, wealth, or urban residence after full adjustment (Unegbu, 2026). The present study addresses the upstream question: what determines whether women achieve this level of ANC in the first place?

The specific objectives were:

1. to estimate the weighted national prevalence of adequate ANC attendance with 95% confidence intervals;
2. to identify independent sociodemographic predictors of adequate ANC after adjustment for confounders;
3. to quantify the extent of confounding in crude associations;
4. to estimate the median gestational age at first ANC visit nationally and by subgroup; and
5. to identify independent predictors of earlier ANC initiation using Cox proportional hazards modelling.

## 2. Methods

### 2.1 Study Design and Data Source

This was a cross-sectional analysis of secondary data from the 2018 Nigeria Demographic and Health Survey (NDHS), a nationally representative household survey conducted by the National Population Commission (NPC) with technical support from ICF under the DHS Program. The survey used a stratified two-stage cluster sampling design. In the first stage, enumeration areas (clusters) were selected with probability proportional to size. In the second stage, households were systematically sampled within each cluster. Women aged 15-49 years residing in selected households were eligible for the individual women’s questionnaire. Full methodological details are reported in the official survey report (NPC & ICF, 2019). DHS data are de-identified and publicly available upon registration at dhsprogram.com. No additional ethical approval was required for this secondary analysis, as the study used only openly available, de-identified data collected under the DHS Program’s standard ethical procedures.

### 2.2 Study Population and Analytic Sample

The individual recode file (NGIR7BFL.DTA) contained records for 41,821 women aged 15-49. The analytic sample was restricted to women who reported at least one live birth in the five years preceding the survey (n = 21,792). Women with missing data on ANC visit count (n = 327, 1.5%) were excluded, yielding a final logistic regression sample of 21,465 women. For the survival analysis examining time to first ANC visit (DHS variable m13_1), a subset of 16,084 women with complete ANC timing data was used. Women excluded from the survival analysis due to missing timing data may differ systematically from those included, which represents a potential source of selection bias and is acknowledged as a study limitation.

### 2.3 Outcome Variables

#### Logistic regression outcome

Adequate ANC attendance was defined as attending four or more ANC visits during the most recent pregnancy, consistent with the WHO 2002 minimum standard as recorded in the 2018 NDHS (DHS variable m14_1). This was coded as a binary variable: 1 = adequate (4 or more visits), 0 = inadequate (fewer than 4 visits).

#### Survival analysis outcome

Time to first ANC visit was defined as the gestational age in months at which the woman attended her first antenatal care visit (DHS variable m13_1). The event of interest was attendance at a first ANC visit. Women who reported never attending ANC were treated as censored at the end of the gestational period.

### 2.4 Explanatory Variables

Seven explanatory variables were included based on a priori theoretical and empirical grounds:

- **Maternal education** (v106): no education (reference), primary, secondary, higher
- **Household wealth index** (v190): poorest (reference), poorer, middle, richer, richest
- **Place of residence** (v025): rural (reference), urban
- **Geopolitical region** (v024): North West (reference), North Central, North East, South East, South South, South West
- **Maternal age** (v012): 15-19 (reference), 20-24, 25-29, 30-34, 35-39, 40-49
- **Parity** (v201): 1 child (reference), 2-3, 4-5, 6 or more
- **Sampling weight** (v005): applied as v005/1,000,000 throughout all analyses

### 2.5 Statistical Analysis

#### Survey design

A complex survey design object was constructed using the srvyr package, specifying cluster IDs (v001) as primary sampling units, strata (v023; 74 state-by-urban/rural strata), and sampling weights. Clusters were nested within strata. All logistic regression estimates account for this design and produce nationally representative estimates.

#### Descriptive analysis

Unweighted sample characteristics were tabulated by ANC adequacy status. Weighted prevalence of adequate ANC was calculated overall and stratified by each explanatory variable, with 95% confidence intervals derived from the survey design.

#### Logistic regression

Unadjusted odds ratios were estimated from simple survey-weighted logistic regression models for each predictor individually using svyglm() with a quasibinomial family. Adjusted odds ratios (aOR) were estimated from a single multivariable model including all seven predictors simultaneously. Odds ratios were exponentiated from log-odds coefficients and 95% confidence intervals derived accordingly. P-values were obtained from Wald tests with a significance threshold of alpha = 0.05.

#### Confounding assessment

Percentage change from crude to adjusted OR was calculated as ((aOR - crude OR) / crude OR) x 100 for each key predictor.

#### Survival analysis

A survival object was constructed using the Surv() function with time defined as months pregnant at first ANC visit. Kaplan-Meier curves were estimated overall and stratified by education, wealth, residence, and region using survfit(), and plotted using survminer::ggsurvplot(). Log-rank tests were used to formally test differences between groups. A Cox proportional hazards model was fitted with all predictors entered simultaneously and sampling weights applied. Hazard ratios greater than 1 indicate faster ANC initiation (earlier attendance); hazard ratios less than 1 indicate slower initiation (later attendance).

All analyses were conducted in R version 4.5.3 using the tidyverse, haven, survey, srvyr, survival, survminer, gtsummary, and flextable packages.

## 3. Results

### 3.1 Sample Characteristics

The logistic regression analytic sample comprised 21,465 women with a birth in the five years preceding the 2018 NDHS. The majority resided in rural areas (64.6%), and the largest education category was no education (44.4%). The North West zone contributed the greatest proportion of the sample (29.4%). The survival analysis sample comprised 16,084 women with complete ANC timing data. Sample characteristics are presented in Table 1.

**Table 1.**
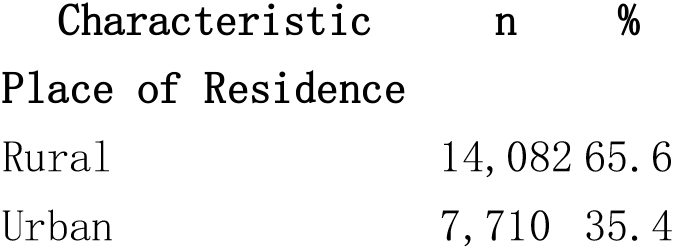
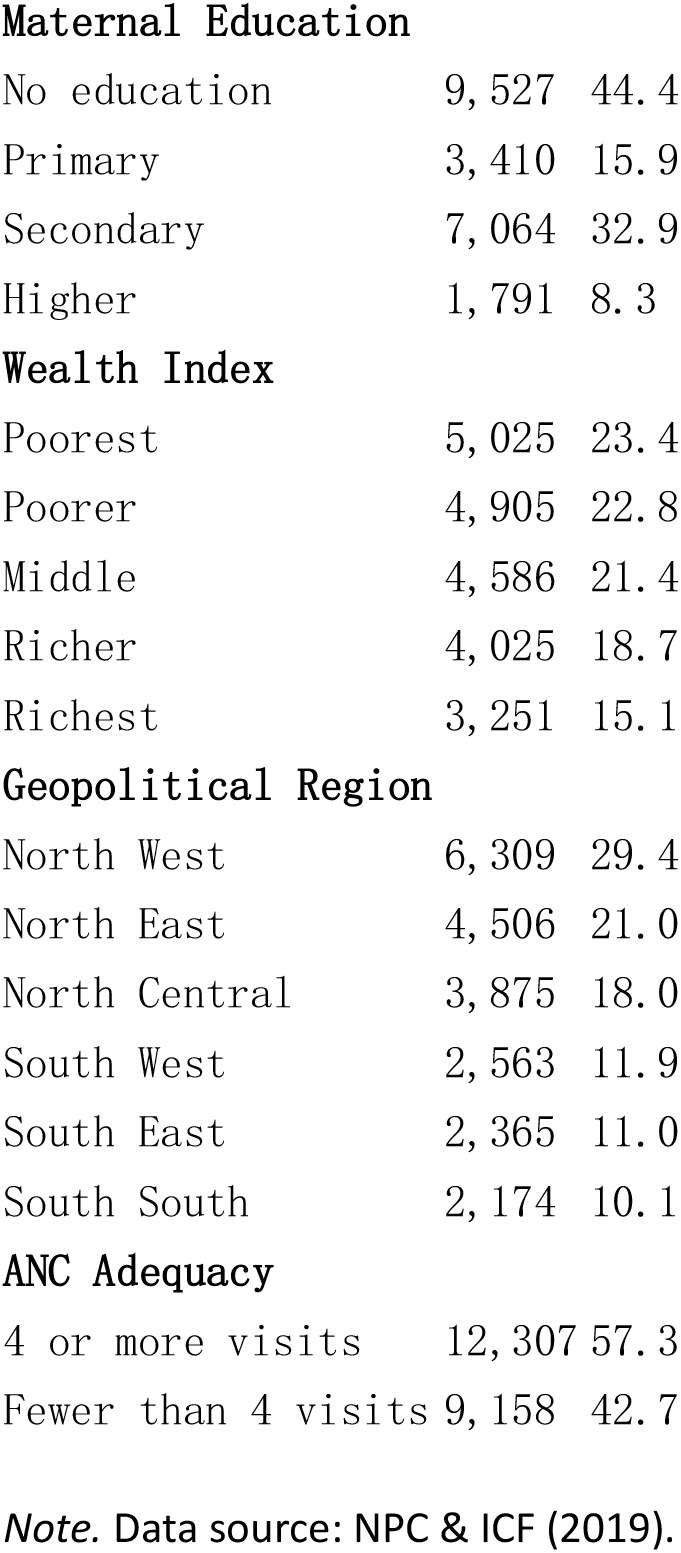
Sociodemographic characteristics of the analytic sample (n = 21,465)

| Characteristic | n | % |
| --- | --- | --- |
| Place of Residence |  |  |
| Rural | 14,082 | 65.6 |
| Urban | 7,710 | 35.4 |
| **Maternal Education** |  |  |
| No education | 9,527 | 44.4 |
| Primary | 3,410 | 15.9 |
| Secondary | 7,064 | 32.9 |
| Higher | 1,791 | 8.3 |
| **Wealth Index** |  |  |
| Poorest | 5,025 | 23.4 |
| Poorer | 4,905 | 22.8 |
| Middle | 4,586 | 21.4 |
| Richer | 4,025 | 18.7 |
| Richest | 3,251 | 15.1 |
| **Geopolitical Region** |  |  |
| North West | 6,309 | 29.4 |
| North East | 4,506 | 21.0 |
| North Central | 3,875 | 18.0 |
| South West | 2,563 | 11.9 |
| South East | 2,365 | 11.0 |
| South South | 2,174 | 10.1 |
| **ANC Adequacy** |  |  |
| 4 or more visits | 12,307 | 57.3 |
| Fewer than 4 visits | 9,158 | 42.7 |

*Note.* Data source: NPC & ICF (2019).

### 3.2 Weighted Prevalence of Adequate ANC Attendance

The national weighted prevalence of adequate ANC attendance was 57.8% (95% CI: 56.2%-59.4%), indicating that approximately 4 in 10 Nigerian women with a recent birth did not meet the WHO minimum standard of four visits. Substantial variation was observed across all sociodemographic subgroups (Table 2). A clear dose-response gradient was observed for both education (34.6% for no education to 92.3% for higher education) and wealth (30.7% for the poorest quintile to 89.2% for the richest). The crude urban-rural gap was marked (76.1% vs. 46.0%), though this attenuated substantially after adjustment (see Section 3.3). Regional disparities were pronounced: adequate ANC prevalence ranged from 42.3% in the North West to 89.9% in the South West, a nearly 48 percentage point gap within a single country.

**Table 2.** Weighted prevalence of adequate ANC attendance by sociodemographic characteristics.

| Variable | Weighted % | 95% CI |
| --- | --- | --- |
| **Overall** | 57.8 | 56.2–59.4 |
| **Education** |  |  |
| No education | 34.6 | 32.5–36.7 |
| Primary | 64.2 | 61.6–66.7 |
| Secondary | 78.6 | 77.3–79.9 |
| Higher | 92.3 | 90.8–93.7 |
| **Residence** |  |  |
| Rural | 46.0 | 44.0–48.0 |
| Urban | 76.1 | 74.2–77.9 |
| **Wealth Index** |  |  |
| Poorest | 30.7 | 28.0–33.4 |
| Poorer | 42.9 | 40.1–45.6 |
| Middle | 61.3 | 59.1–63.6 |
| Richer | 75.3 | 73.2–77.3 |
| Richest | 89.2 | 87.7–90.7 |
| **Region** |  |  |
| North West | 42.3 | 39.4–45.2 |
| North East | 44.0 | 40.8–47.3 |
| North Central | 54.6 | 51.6–57.6 |
| South South | 72.6 | 69.5–75.6 |
| South East | 84.3 | 82.2–86.3 |
| South West | 89.9 | 87.8–92.0 |

### 3.3 Multivariable Logistic Regression

Results from the fully adjusted model are presented in Table 3. Higher education was the strongest independent predictor of adequate ANC (aOR = 5.64, 95% CI: 4.45-7.15, p < .001), followed by the richest wealth quintile (aOR = 3.93, 95% CI: 3.11-4.95, p < .001). A monotonic dose-response gradient was observed for both education and wealth, with each successive category associated with progressively higher odds of adequate ANC attendance.

**Table 3.**
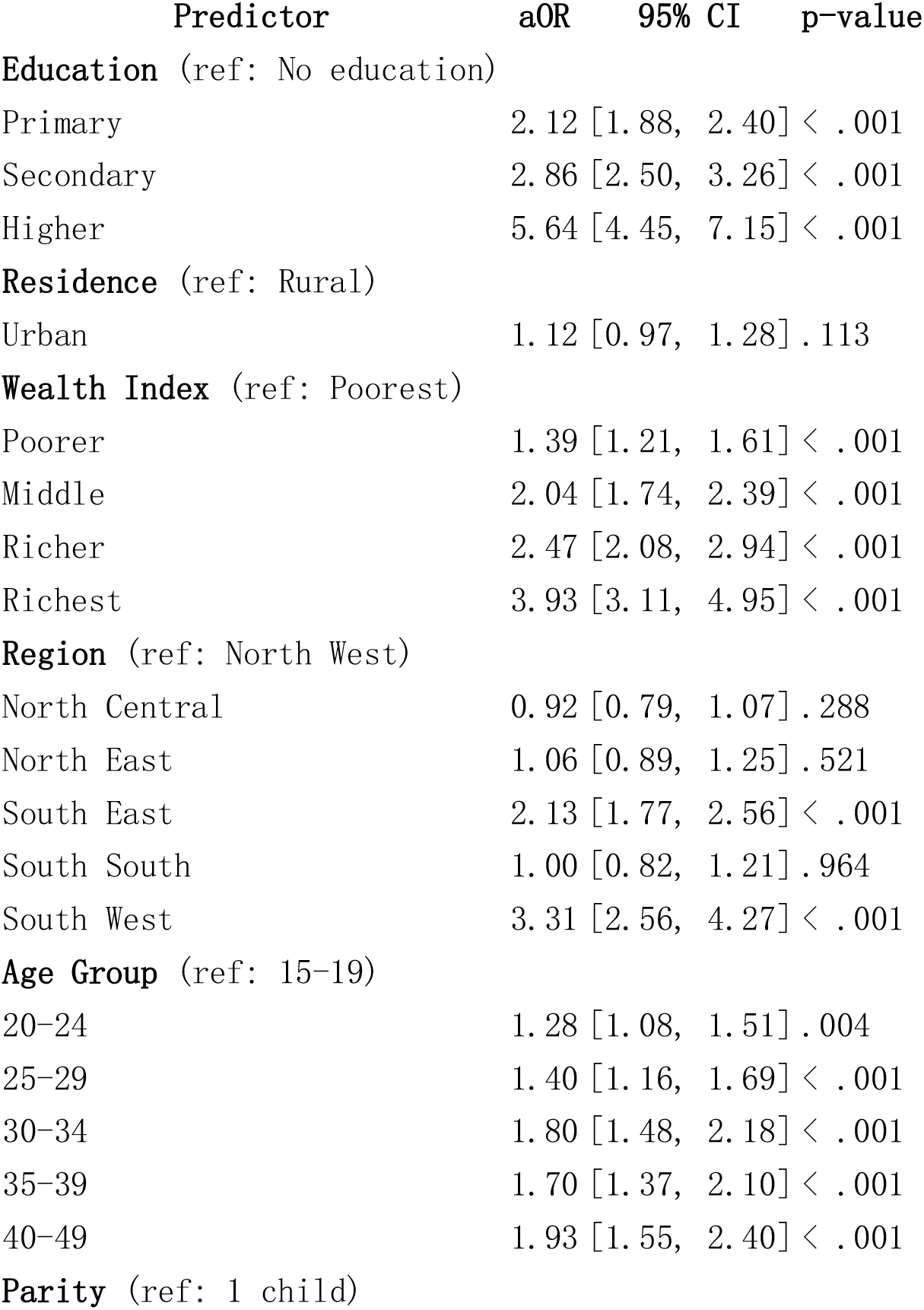
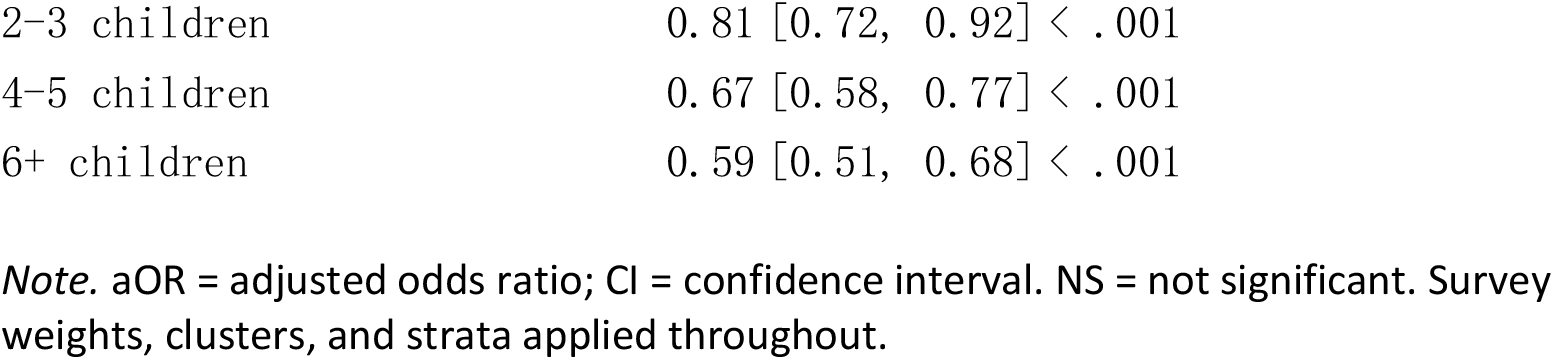
Survey-weighted multivariable logistic regression: adjusted odds ratios for adequate ANC attendance (n = 21,465)

The most notable finding in the adjusted model was the complete loss of significance for urban residence (aOR = 1.12, 95% CI: 0.97-1.28, p = .113). Despite the large crude urban-rural gap in ANC prevalence (76.1% vs. 46.0%), urban residence carried no independent protective effect after controlling for education, wealth, and region. This indicates that the apparent urban advantage operates entirely through the higher educational attainment and household wealth of urban women, rather than through any inherent characteristic of urban living or proximity to urban health infrastructure.

Among regional predictors, only the South West (aOR = 3.31, 95% CI: 2.56-4.27) and South East (aOR = 2.13, 95% CI: 1.77-2.56) showed significant independent effects relative to the North West reference category. North Central, North East, and South South regions were not independently significant after adjustment, suggesting their crude differences from the North West are largely mediated by their socioeconomic composition. Higher parity was independently associated with lower odds of adequate ANC attendance in a dose-response pattern, with women with six or more children having 41% lower odds compared to primiparous women (aOR = 0.59, 95% CI: 0.51-0.68, p < .001). Older maternal age was positively associated with adequate ANC from age 20 onwards, with the strongest effect in the 40-49 age group (aOR = 1.93, 95% CI: 1.55-2.40).

### 3.4 Confounding Assessment

Crude odds ratios were substantially larger than adjusted estimates for all education categories (Table 4). The attenuation was most extreme for higher education (crude OR = 22.49 vs. aOR = 5.64; 74.9% reduction), indicating that the majority of its crude association with adequate ANC is attributable to confounding by correlated wealth, residence, and regional factors. Urban residence showed the most dramatic confounding: its large crude effect was entirely explained by adjustment, yielding a non-significant adjusted estimate. This pattern — the complete disappearance of the urban effect after adjustment — is a substantively important finding that challenges the common policy assumption that urbanization per se improves maternal health service utilization.

**Table 4.** Crude versus adjusted odds ratios: magnitude of confounding.

| Variable | Crude OR | Adjusted OR | % Change | Interpretation |
| --- | --- | --- | --- | --- |
| Higher education | 22.49 | 5.64 | –74.9% | Heavily confounded by wealth and residence |
| Secondary education | 6.94 | 2.86 | –58.9% | Substantial confounding |
| Primary education | 3.39 | 2.12 | –37.3% | Moderate confounding |
| Urban residence | Large | 1.12 (NS) | Lost significance | Effect fully explained by wealth and education |

### 3.5 Survival Analysis: Time to First ANC Visit

#### 3.5.1 Kaplan-Meier Analysis

The national median gestational age at first ANC visit was 5 months (95% CI: 4-5 months), two months later than the WHO recommendation of initiating care before the end of the third month of pregnancy (WHO, 2002). The Kaplan-Meier table illustrates the distribution of ANC initiation across gestational months (Table 5).

**Table 5.** Kaplan-Meier estimates: proportion of women not yet attending first ANC visit by gestational month.

Month of Pregnancy % Not Yet Attended First ANC
|  |  |
| --- | --- |
| Month 1 | 98.0% |
| Month 2 | 92.6% |
| Month 3 | 75.9% |
| Month 4 | 50.1% |
| Month 5 | 27.7% |
| Month 6 | 12.6% |
| Month 7 | 2.8% |

The peak period of ANC initiation was months 3 to 5 of pregnancy, with the largest single-month drop occurring between months 4 and 5. By month 7, only 2.8% of women had not yet attended their first visit. Log-rank tests confirmed significant differences in ANC initiation timing across all sociodemographic subgroups (all p < .0001; Table 6).

**Table 6.**
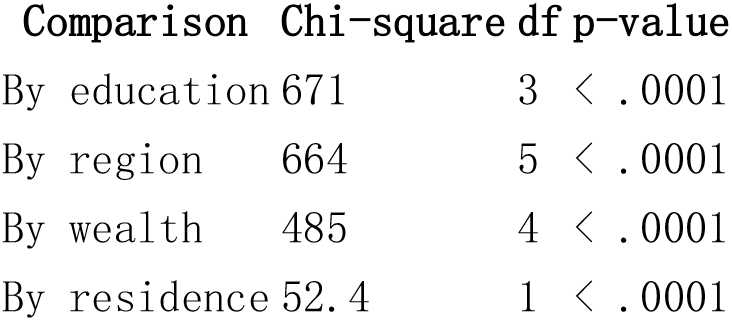
Log-rank test results for differences in ANC initiation timing by subgroup.

Kaplan-Meier curves by education showed clear separation across all four groups, with higher educated women exhibiting the steepest early decline indicating the fastest ANC initiation. Uneducated women remained at the highest survival proportion longest, reflecting the latest initiation. By region, the north-south divide in ANC timing was visually striking: at month 5, 2,493 North West women had still not attended their first visit, compared with 906 South West women a 2.7-fold difference.

#### 3.5.2 Cox Proportional Hazards Model

Results from the weighted Cox proportional hazards model are presented in Table 7. Higher education was the strongest predictor of earlier ANC initiation (HR = 1.35, 95% CI: 1.23-1.47, p < .001), indicating that women with higher education initiated ANC 35% faster than uneducated women after adjustment. A dose-response pattern was observed across education levels. Wealth also independently predicted earlier initiation, with richest quintile women initiating 32% faster than the poorest (HR = 1.32, 95% CI: 1.22-1.43).

**Table 7.** Weighted Cox proportional hazards model: hazard ratios for time to first ANC visit (n = 16,084)

| Predictor | HR | 95% CI | p-value |
| --- | --- | --- | --- |
| **Education** (ref: No education) |  |  |  |
| Primary | 1.10 | [1.04, 1.16] | < .001 |
| Secondary | 1.19 | [1.13, 1.26] | < .001 |
| Higher | 1.35 | [1.23, 1.47] | < .001 |
| **Residence** (ref: Rural) |  |  |  |
| Urban | 0.93 | [0.89, 0.97] | < .001 |
| **Wealth Index** (ref: Poorest) |  |  |  |
| Poorer | 1.07 | [1.02, 1.13] | .011 |
| Middle | 1.11 | [1.05, 1.17] | < .001 |
| Richer | 1.14 | [1.07, 1.22] | < .001 |
| Richest | 1.32 | [1.22, 1.43] | < .001 |
| **Region** (ref: North West) |  |  |  |
| North Central | 1.29 | [1.22, 1.37] | < .001 |
| North East | 1.13 | [1.08, 1.19] | < .001 |
| South East | 1.22 | [1.14, 1.30] | < .001 |
| South South | 1.13 | [1.06, 1.22] | < .001 |
| South West | 1.36 | [1.26, 1.46] | < .001 |
| **Parity** (ref: 1 child) |  |  |  |
| 4-5 children | 0.80 | [0.74, 0.87] | < .001 |
| 6+ children | 0.77 | [0.71, 0.84] | < .001 |

*Note.* HR = hazard ratio; CI = confidence interval. HR > 1 indicates faster ANC initiation (earlier attendance); HR < 1 indicates slower initiation. Survey weights applied. Full complex survey variance estimation (clusters and strata) was not implemented for the Cox model; standard errors may be slightly underestimated.

The residence finding in the Cox model reinforced the logistic regression result, but with an additional nuance: urban women actually initiated ANC more slowly than rural women after adjustment (HR = 0.93, 95% CI: 0.89-0.97, p < .001). This counterintuitive finding suggests that after removing the confounding influence of education and wealth, rural women initiate care at a similar or slightly faster pace — possibly reflecting community health worker outreach in rural areas. All regional zones initiated ANC significantly earlier than the North West reference, with the South West showing the fastest initiation (HR = 1.36, 95% CI: 1.26-1.46). Higher parity was independently associated with slower initiation (HR = 0.77 for 6 or more children, 95% CI: 0.71-0.84, p < .001).

## 4. Discussion

This nationally representative analysis of 21,465 Nigerian women provides a comprehensive picture of the determinants of adequate ANC attendance and ANC initiation timing, using both logistic regression and survival analysis approaches. The national weighted prevalence of adequate ANC was 57.8%, confirming that despite modest progress, 4 in 10 Nigerian women continue to fall below the WHO minimum standard of four visits. The median gestational age at first ANC visit of 5 months — two months later than WHO recommendations — is a clinically significant finding that compounds the inadequacy of total visit counts: women who initiate late have less time to accumulate sufficient visits and less time to benefit from the interventions ANC provides.

The finding that higher education was the strongest independent predictor of adequate ANC attendance (aOR = 5.64) is consistent with a broad literature linking maternal education to maternal health service utilization in sub-Saharan Africa through pathways of health literacy, autonomous decision-making, and reduced financial dependence (Fagbamigbe & Idemudia, 2015; Tessema, Z. T., et. al 2021). The survival analysis extended this finding temporally: educated women did not merely attend more visits, they initiated care 35% faster than uneducated women (HR = 1.35), compressing the gestational window over which interventions are delivered. This dual effect — on quantity and timing of care — makes girls’ education the single most upstream and highest-leverage intervention available for improving maternal health outcomes in Nigeria.

The complete loss of significance for urban residence after adjustment (aOR = 1.12, p = .113) is one of the most substantively important findings of this analysis. The crude prevalence gap between urban and rural women was 30 percentage points (76.1% vs. 46.0%), a difference that would, if taken at face value, strongly support urban-focused health investment. After controlling for the higher education and wealth concentrated in urban areas, this gap disappears entirely. The Cox model reinforced this with an additional counterintuitive result: after adjustment, urban women initiated ANC slightly more slowly than rural women (HR = 0.93). This pattern is consistent with findings from comparable DHS analyses in other West African settings and challenges the assumption that urbanization per se drives maternal health behavior (Moyer & Mustafa, 2013). The policy implication is direct: investment in urban health infrastructure will not close the ANC gap unless it is accompanied by investment in the education and economic empowerment of women in those settings.

The persistence of a strong independent wealth effect (aOR = 3.93 for richest vs. poorest) after adjustment for education and residence confirms that economic barriers to ANC operate through mechanisms beyond geography or educational attainment. With adequate ANC coverage of only 30.7% in the poorest quintile, direct financial barriers — transport costs, user fees at referral facilities, and opportunity costs of attending clinic — remain structural obstacles for the poorest women. Free-at-point-of-use ANC policies address only one dimension of this barrier; demand-side interventions including conditional cash transfers, targeted transport subsidies, and community-based outreach are likely needed to shift behavior in the poorest quintile.

The pronounced regional gradient in both ANC attendance and initiation timing, persisting after adjustment, reflects supply-side differences in health infrastructure density, health worker availability, and socio-cultural norms that are not captured by individual-level socioeconomic variables. The North West and North East — where adjusted ANC odds were lowest and initiation was latest — require the most intensive programmatic investment. Community health worker home-visiting programmes targeting pregnant women in months 2 and 3 of pregnancy in these regions offer a directly actionable, evidence-based entry point.

The inverse parity-ANC relationship observed in the logistic regression (aOR = 0.59 for 6 or more children) is consistent with findings from comparable analyses in sub-Saharan Africa and reflects the well-documented tendency for high-parity women to perceive lower risk in subsequent pregnancies based on prior uncomplicated deliveries (Mpembeni et al., 2007). The Cox model showed that high-parity women also initiated ANC later (HR = 0.77 for 6 or more children), confirming that this group is disadvantaged on both dimensions of care quality. Targeted behaviour change communication addressing risk normalization in high-parity women is warranted.

This paper forms the second study in a two-paper evidence chain using the same 2018 NDHS dataset. The companion SBA analysis established that attending four or more ANC visits was the strongest modifiable predictor of skilled birth attendance (aOR = 3.80) (Unegbu, 2026). The present analysis establishes that education and wealth are the dominant upstream drivers of achieving that level of ANC. Together, the two studies trace the causal pathway from structural socioeconomic factors through ANC utilization to skilled delivery, providing a coherent evidence base for maternal health programme design in Nigeria.

### 4.1 Strengths and Limitations

Key strengths include the use of a large, nationally representative dataset with full accounting for complex survey design, the application of two complementary analytic methods capturing both whether and when women access ANC, and the explicit quantification of confounding. The survival analysis dimension is rarely applied in the Nigerian ANC literature and adds clinically relevant information about care timing that aggregate utilization statistics do not capture.

Several limitations warrant acknowledgment. First, the cross-sectional design precludes causal inference. Second, ANC attendance was self-reported and subject to recall bias, as women reported on births up to five years prior. Third, the survival analysis sample (n = 16,084) was smaller than the logistic regression sample (n = 21,465) due to missing ANC timing data, and women excluded may differ systematically from those included. Fourth, the Cox model incorporated survey weights but not full complex survey variance estimation using clusters and strata; standard errors in the Cox results may therefore be slightly underestimated. Fifth, important potential confounders including distance to the nearest health facility, quality of care, and women’s autonomous decision-making power were not available in the dataset. Finally, the 2018 data predate the COVID-19 pandemic, which significantly disrupted maternal health services globally and may have altered ANC attendance patterns in Nigeria post-2020.

## 5. Conclusions

Education and wealth are the dominant independent determinants of both adequate ANC attendance and earlier ANC initiation in Nigeria. The large crude urban-rural difference in ANC coverage is entirely explained by the higher education and wealth of urban women, with no independent urban effect remaining after adjustment — a finding with direct implications for how ANC investment is geographically targeted. The median gestational age at first ANC visit of 5 months, two months later than WHO recommendations, compounds inadequate visit counts with insufficient time to benefit from antenatal interventions. Urgent investment in girls’ education, wealth-sensitive demand-side financing, and community-based early ANC outreach — with particular intensity in the North West and North East — is needed to close Nigeria’s ANC gap and, through it, reduce the country’s unacceptably high burden of preventable maternal mortality.

## Data Availability

The data used in this study are publicly available from the DHS Program upon free registration. The 2018 Nigeria Demographic and Health Survey individual recode dataset (NGIR7BFL.DTA) can be accessed at: https://dhsprogram.com/data/dataset/Nigeria_Standard-DHS_2018.cfm.

